# *SwissGenVar*: A platform for clinical grade interpretation of genetic variants to foster personalized health care in Switzerland

**DOI:** 10.1101/2023.01.11.22283790

**Authors:** Dennis Kraemer, Dillenn Terumalai, Maria Livia Famiglietti, Isabel Filges, Pascal Joset, Samuel Koller, Fabienne Maurer, Stéphanie Meier, Thierry Nouspikel, Javier Sanz, Christiane Zweier, Marc Abramowicz, Wolfgang Berger, Sven Cichon, André Schaller, Andrea Superti-Furga, Valérie Barbié, Anita Rauch

**Affiliations:** Institute of Medical Genetics (IMG), University of Zurich (UZH), CH-8952 Schlieren/Zurich, Switzerland; Swiss Institute of Bioinformatics (SIB), Clinical Bioinformatics, Campus Biotech, CH-1202 Geneva, Switzerland; Swiss Institute of Bioinformatics (SIB), Swiss-Prot group, Centre Medicale Universitaire (CMU), CH-1211 Geneva, Switzerland; Medical Genetics, Institute of Medical Genetics and Pathology, University Hospital Basel and University of Basel, CH-4031 Basel, Switzerland; Institute of Medical Molecular Genetics (IMMG), University of Zurich (UZH), CH-8952 Schlieren/Zurich, Switzerland; Division of Genetic Medicine, Lausanne University Hospital (CHUV), CH-1011 Lausanne, Switzerland; Genetic Medicine Division, Diagnostics Dept./Center for Genomic Medicine, Geneva University Hospitals (HUG), Geneva, Switzerland; Department of Human Genetics, Inselspital, Bern University Hospital, CH-3010 Bern, Switzerland; Neuroscience Center Zurich (ZNZ), University and ETH Zurich, Zurich, Switzerland; Zurich Center for Integrative Human Physiology (ZIHP), University of Zurich, Zurich, Switzerland

**Keywords:** *SwissGenVar*, Switzerland, NGS, expert-curated variant interpretation, national mutation database, genotype-phenotype database, personalized medicine

## Abstract

Large-scale next-generation sequencing (NGS) germline testing is technically feasible today, but variant interpretation represents a major bottleneck in analysis workflows including the extensive variant prioritization, annotation, and time-consuming evidence curation. The scale of the interpretation problem is massive, and variants of uncertain significance (VUS) are a challenge to personalized medicine. This challenge is further compounded by the complexity and heterogeneity of standards used to describe genetic variants and associated phenotypes when searching for relevant information to inform clinical decision-making.

For this purpose, all five Swiss academic Medical Genetics Institutions joined forces with the Swiss Institute of Bioinformatics (SIB) to create *SwissGenVar* as a user-friendly nationwide repository and sharing platform for genetic variant data generated during routine diagnostic procedures and research sequencing projects. Its objective is to provide a protected environment for expert evidence sharing about individual variants to harmonize and up-scale their significance interpretation at clinical grade following international standards. To corroborate the clinical assessment, the variant-related data are combined with consented high-quality clinical information. Broader visibility will be gained by interfacing with international databases, thus supporting global initiatives in personalized health care.

## 1. Introduction

Assessment of individual genetic risk factors and classification of molecular disease based on genetic contributions are hallmarks of personalized medicine [1-5]. Next to common genetic variants predisposing to, or modulating common diseases, newer evidence also indicates a significant role of individually rare variants in frequently mutated genes with strong functional consequences [6, 7]. Large-scale germline genetic testing is technically feasible today but is hampered by the difficulties in interpreting the clinical significance of variants, lack of knowledge about genotype-phenotype correlation, and long-term clinical history [8, 9]. Accurate pathogenicity interpretation of genetic variants is not only crucial for appropriate medical decision-making based on genetic evidence [10] but also for the correct stratification of research findings by genetic results [11]. A variety of international database initiatives aim to facilitate genetic variant assessment; however, these often are restricted to specific genes and/or (types of) genetic diseases/alterations and contain insufficient or contradictory, sometimes even incorrect public entries [12, 13], which mostly fail to provide accompanying valid clinical data for variant interpretation in their respective phenotypic contexts [14, 15]. Moreover, since genetic variation commonly differs among ethnicities, international data collection may not be representative and comprehensive for specific populations. The importance of local and national genetics has been exemplified by the Genome of the Netherlands initiative [16]. Therefore, the next big challenge in personalized medicine will be the expansion of high-quality genotype-phenotype databases providing “knowledge” over “data” to enable, without dictating, accurate clinical care due to rigid quality management and a sustainable expert variant curation and classification process [17, 18].

In Switzerland, research including human genetic data, as well as diagnostic germline genetic testing is strictly regulated and subject to regular quality control and accreditation procedures. Accordingly, research involving germline genetic data and diagnostic genetic testing is mainly pursued by highly specialized centers including the university and cooperating clinical centers for Medical/Human Genetics. The use of next-generation sequencing (NGS) technologies as the standard of care creates a rich source of genetic data with an in-depth clinical variant assessment that currently is not collected systematically. In Switzerland so far, there exists no nationwide academic/public database for genetic variants obtained from diagnostic procedures or sequencing research projects. Therefore, the considerable potential to promote sharing diagnostic-grade genomic data with patient-related consented high-quality clinical information remains largely untapped. Besides data protection issues, this may be explained, particularly for Swiss institutions, by the previous lack of agreed harmonized standards and concepts for genetic data collection and exchange as well as the absence of a suitable and secure repository infrastructure for genomic and related patient data.

To address these difficulties and leverage the high-quality genetic and accompanying clinical data generated in Swiss academic institutions, all five Swiss university centers for Medical/Human Genetics joined forces with the Swiss Institute of Bioinformatics (SIB, Clinical Bioinformatics) in a nationwide effort to create the *SwissGenVar* platform within the framework of a Swiss Personalized Health Network (SPHN) [19] infrastructure development project (project page available at [20]). *SwissGenVar* aims at providing a protected nationwide repository for germline variants identified in patients by Swiss clinical genetic laboratories with accompanying high-quality clinical data and an efficient joint platform for harmonization and up-scaling of expert-curated variant interpretation. However, *SwissGenVar* not only fosters harmonization and inclusion of diagnostic data, but also of data generated within research projects using genomic sequencing approaches. To this end, *SwissGenVar* ensures interoperability with international databases and the methodological and technical prerequisites for national and international sharing/storage of genomic data and evidence for standardized variant pathogenicity assessment, to facilitate consensus variant classification by clinical genetic experts. Furthermore, *SwissGenVar* allows the harvesting of patient-consented clinical data generated during routine health care to assess the clinical significance of a variant for a specific disease in synopsis with associated phenotypic features.

Within this project, we, therefore, defined an interoperable set of genetic and non-identifying clinical data for variant data sharing/storage and clinical interpretation, a consistent genetic variant file upload and annotation process as well as a data ontology appropriate for the creation of a protected nationwide germline variant database with accompanying high-quality clinical data and significance interpretation. The accessibility for clinicians and researchers has been realized through an efficient, scalable, and user-friendly IT infrastructure, integrated within the secure BioMedIT [21] landscape of SPHN. Currently, *SwissGenVar* is accessible to the project partners only, but with a scope for expansion to further academic and non-academic institutions to establish itself as the Swiss one-stop platform for the interpretation/understanding of genetic germline variants. Thus, *SwissGenVar* may substantially foster personalized health care, and, on the other hand, be a necessary first step to scale-up clinical-grade genetic testing and data sharing in Switzerland.

## 2. Materials and Methods

### 2.1 Sensitive data hosting and transfers

The *SwissGenVar* infrastructure development project has been initially funded by the Swiss Personalized Health Network (SPHN) [19] initiative which builds on the Swiss national BioMedIT infrastructure, specifically implemented for hosting sensitive data. It, therefore, uses all tools provided by those initiatives and follows their requirements.

The *SwissGenVar* application and data are hosted on the secure SENSA (Secure Sensitive Data Processing Platform) BioMedIT [21] node in Lausanne and comply with the SPHN and BioMedIT tools and the related Information Security Policy [22]. Data transfers are ensured by the SPHN SETT (Secure Encryption and Transfer Tool) data transfer tool [23], which encrypts, securely transfers, and decrypts data.

Users’ identity and access are managed using the BioMedIT central Keycloak [24] instance, which requires SWITCH eduID [25] two-factor authentication to log in. Keycloak is an open-access IAM platform that secures web applications and RESTful web services using standard protocols such as OAuth2, OpenID Link, and SAML 2.0. In addition, access to the system is restricted to the whitelisted IP (Internet Protocol) address ranges of each participating institution.

All data used for the development of the platform and depicted in the figures are for fictitious individuals, not real patients.

### 2.2 Software development

*SwissGenVar* is a web-based application, which backend is written in PHP (using the Laravel framework) and relies on a PostgreSQL database. The frontend is based on Vue.js (using the Nuxt framework). The bioinformatics pipeline is running on a SLURM cluster.

### 2.3 Public data sources

For all variants in the VCF (Variant Call Format) files, some public information is automatically gathered by the *SwissGenVar* platform using a local instance of the Ensembl Variant Effect Predictor (VEP) [26] deployed on the SENSA BioMedIT node. This information currently includes the variant type and effect, the genomic position of the variant, and the HGVS (Human Genome Variation Society) variant nomenclature [27].

## 3. Results

### 3.1 *SwissGenVar* governance and layers of access

For implementation and administration of *SwissGenVar*, a multicenter consortium among all five academic centers for Medical Genetics in Switzerland and the Swiss Institute of Bioinformatics (SIB) (Figure 1) has been formed, which is governed by the Steering Board, as defined in the *SwissGenVar* Consortium Agreement. To combine the use for research and the highest level of data protection, the platform is composed of two different modules with different potential layers of access, which are specified by the Data Transfer and Use Agreement (DTUA). The access-controlled instance is intended to share genetic data and non-identifying associated clinical/demographic metadata in view-only mode, including data submitted by any other registered group. Registered users belonging to a registered group may in addition modify their own data or metadata. The access to the data stored in the access-controlled instance is restricted to registered users of the consortium (full access layer). However, upon approval by the Steering Board, data from the access-controlled instance (including personal data) may be made accessible to users belonging to a third-party group if required for a specific research study and if authorized by the competent ethics committee (restricted access layer). By contrast, the public instance aims to make stand-alone variants and aggregated patient/proband data (without any information related to the specific patient/proband or sample) publicly available and will be freely accessible by all interested parties without registration (public access layer).

**Figure 1.**
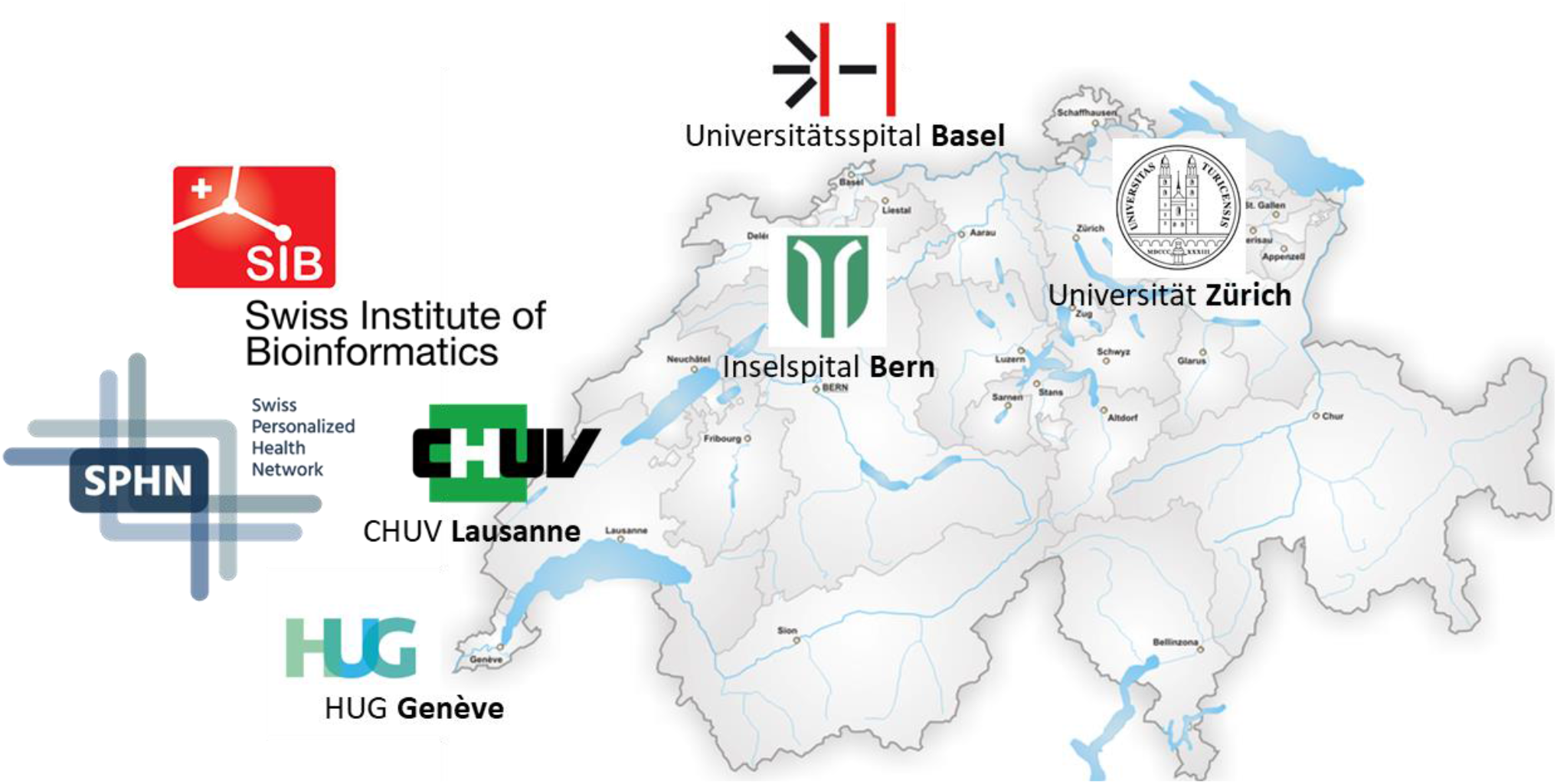
Involved institutions of the *SwissGenVar* consortium: University Hospital Basel, Medical Genetics; Department of Human Genetics, Inselspital, Bern University Hospital; Medical Genetics Service/Center for Genetic Medicine, Geneva University Hospital (HUG); Medical Genetics Service/Division of Genetic Medicine, Lausanne University Hospital (CHUV); Institute of Medical Genetics (IMG), University of Zurich (UZH); Institute of Medical Molecular Genetics (IMMG), University of Zurich (UZH); Swiss Institute of Bioinformatics (SIB); Swiss Personalized Health Network (SPHN).

### 3.2 Standardized *SwissGenVar* dataset specifications

One of the key concepts of *SwissGenVar* is the combination of diagnostic-grade genetic variant-related data and accompanying consented high-quality basic clinical information to corroborate their diagnostic utility. To harmonize the variant-related and phenotypic ontologies, a cross-expert working group defined a minimal and an extended genetic and clinical data set pertinent for data sharing/storage and the standardized interpretation of the clinical significance of genetic variants, which have been approved by the *SwissGenVar* board. After several rounds of thorough discussions and board meetings, dedicated clinical and laboratory working groups, which were managed by clinical experts for the addressed issue, elaborated a comprehensive and granular portfolio of parameters and functionalities needed for the objectives of *SwissGenVar*.

To ensure interoperability with international databases and other SPHN projects, *SwissGenVar* follows established international standards and the SPHN guidelines for Interoperability Data Standard and Tool Collection [22], whenever applicable. For most items, well-defined existing ontologies are used (Tab. 1). However, for the data fields relevant to the *SwissGenVar* project where no appropriate data standard was available, the consortium had to define and adapt an internal data catalogue reflecting the consensus between the practices at the different partner institutions.

**Table 1.**
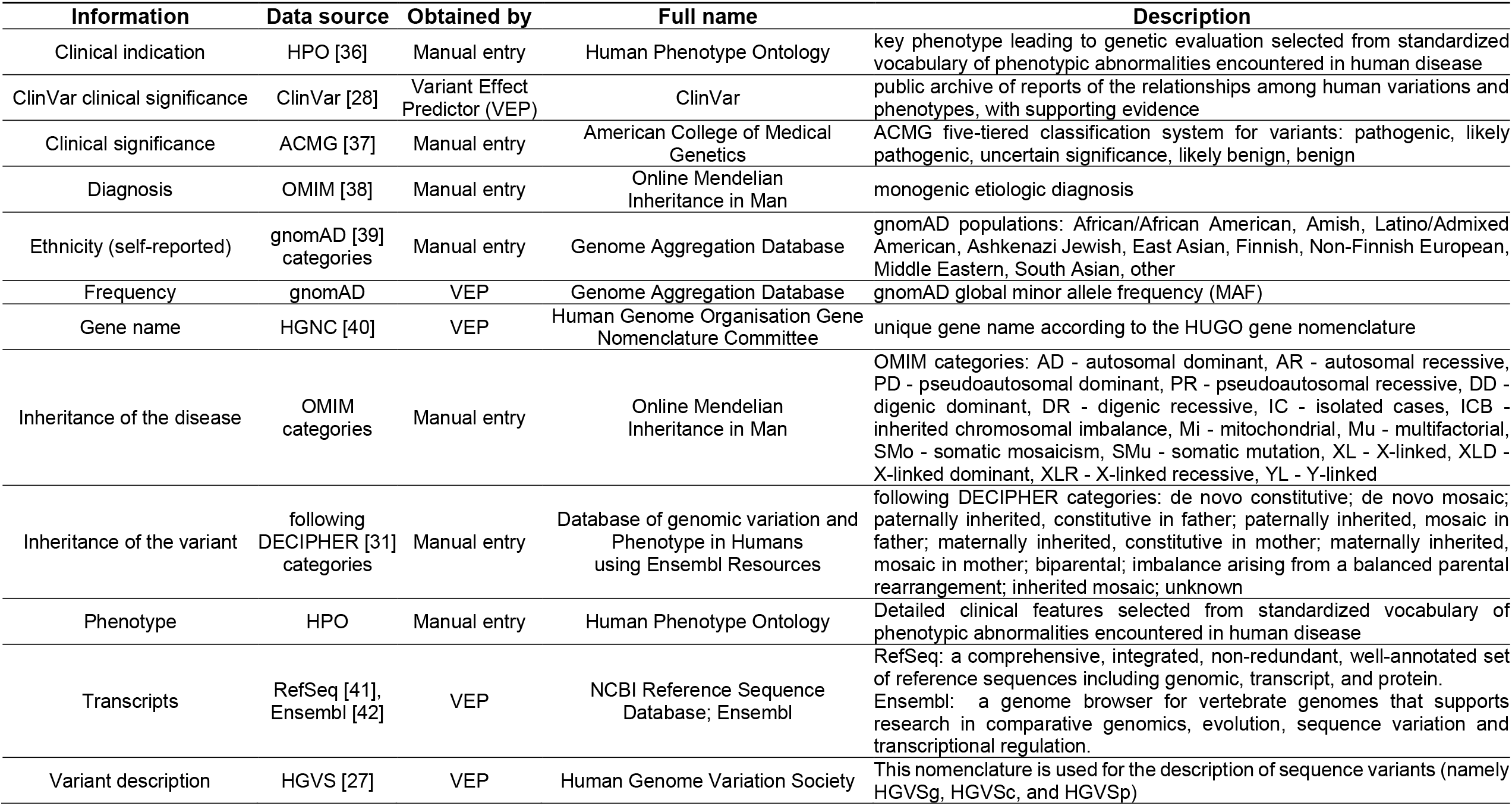
Standardized core dataset. **(A)** Established data catalogues and data sources used in *SwissGenVar*.

**Table 1.** Standardized core dataset. **(B)** Internal data catalogues defined for *SwissGenVar*.

| Information | Possible values | Remark |
| --- | --- | --- |
| Age at onset | -1 (prenatal), 0, 0.1, 0.2, ..., 100 | range of numbers for the age of onset in years |
| Aneuploidies | yes; no |  |
| Canton | list of Swiss cantons, plus "non-Swiss" |  |
| Causality | causative; likely causative; probably not causative; not causative; VUS; variant in a GUS | causality following clinical judgement |
| Chromosomal sex | XX; XY; other |  |
| Clinical gender | male, female, ambiguous, transgender |  |
| Karyotypic sex | 45X, 46XX, 46XY, 47XXY, 47XYY, 47XXX (intended as expandable list) | content is conditional to the value of "other" in "Chromosomal sex" |
| Clinical status | affected; partially affected; potentially affected; not affected | defined fields/filters: "clinical status change to"; "clinical status at last clinical assessment" |
| Co-occurrences | yes; no | co-occurrence of more than one causative variant |
| Collection method | case-control; clinical testing; reference population; research; other; unknown |  |
| Cytogenetic location | the cytogenetic location of the variant displayed as CHROM_NUMBERq/pCYTOGENETIC_BAND |  |
| Detection method | sequencing; fragment analysis; Southern Blot; conventional cytogenetics; FISH (IFISH or MFISH); Array (Oligo or SNP); qRT-PCR; MLPA; NGS-based CNV detection (Panel/WES/WGS); other; not performed |  |
| Gene locus type | protein-coding gene; non-coding RNA gene; long non-coding RNA; microRNA; ribosomal RNA; transfer RNA; small nuclear RNA; small nucleolar RNA; other; locus subjected to imprinting | partly coming from HUGO Gene Nomenclature Committee (HGNC) [40] |
| Index patient | yes; no |  |
| Location | genomic position | GRCh37 as genome reference built [43] |
| Locus subjected to imprinting | yes; no; unknown |  |
| Patient identifier (ID) | the patient ID refers to an internal *SwissGenVar* specific unique identifier that is generated when the patient is created in the system | patient/sample ID of the submitting institution is recorded as well |
| Submitting institution | one acronym per partner institution |  |

**Table 1 (B) continued**
| Information | Possible values | Remark |
| --- | --- | --- |
| Variant effect | missense variant; nonsense variant; splice region variant; splice acceptor variant; splice donor variant; regulatory region variant; promoter region variant; inframe insertion; inframe deletion; intron variant; synonymous variant; stop lost variant; start lost variant; frameshift variant; upstream gene variant; downstream gene variant; intergenic variant; non-coding transcript exon variant; TF binding site variant; 5' UTR variant; 3' UTR variant; exon deletion; exon duplication; contiguous gene deletion; contiguous gene duplication | adapted to Sequence Ontology (SO) [44] terms |
| Variant location | coding region; splicing region; 5' UTR; 3' UTR; upstream gene; downstream gene; promoter region; intronic region; regulatory region; intergenic region |  |
| Variant type | CNV – amplification; CNV – deletion; CNV - insertion/duplication; complex rearrangement; conversion; deletion; deletion-insertion; duplication; insertion; methylation/epigenetic change; repeat variation; structural variant; substitution |  |
| Variant zygosity | heterozygous; homozygous; hemizygous; mitochondrial heteroplasmy; mitochondrial homoplasmy; unknown; mosaic; chimeric; ambiguous |  |

CNV, copy number variation; FISH, fluorescence in situ hybridisation (IFISH, interphase-FISH; MFISH, metaphase-FISH); GUS, gene of uncertain significance; MLPA, multiplex ligation-dependent probe amplification; qRT-PCR, quantitative reverse transcription PCR (polymerase chain reaction); SNP, single nucleotide polymorphism; TF, transcription factor; WES, whole-exome sequencing; WGS, whole-genome sequencing; VUS, variant of unclear significance; UTR, untranslated region

Furthermore, *SwissGenVar* enables automated variant annotation from a variety of sources and implements direct links to the well-established NCBI ClinVar [28] and Single Nucleotide Polymorphism Database (dbSNP) [29] as well as to Human Gene Mutation Database (HGMD) [30], DECIPHER [31], LOVD (Leiden Open Variation Database) [32] and SVIP-O [33], the latter being a Swiss SPHN platform for the clinical interpretation of genetic variants in oncology (Swiss Variant Interpretation Platform for Oncology). Additionally, the widely-used predictive algorithms SIFT (Sorting Intolerant From Tolerant) [34] and PolyPhen-2 [35] for *in silico* assessment of amino acid substitutions are implemented using VEP.

### 3.3 Data management and application workflow

The project partners provide high-quality genetic data mostly from NGS procedures (exome and genome sequencing or other methods in the form of VCF files [Variant Call Format]) either derived from research studies or diagnostic testing with general or dedicated *SwissGenVar* consent, which are complemented by a minimal set of basic clinical information from the relevant medical history of the patient (Figure 2). These genetic and clinical data are generated either directly by the involved laboratories or by the hospital Clinical Data Warehouses (CDW), depending on each partner institution’s setup. In both cases, genetic data are encrypted and securely transferred using the SPHN BioMedIT transfer tool [23] and are stored and accessed according to BioMedIT access and security standards [24, 25]. Subsequently, after decryption and parsing of the transferred files, patient entries are created and variant calls from the VCF files are loaded into the platform. Before being loaded, the genetic data are going through a technical basic check-up to ensure compliance with the requested VCF file format. The user can then select individual variants as “of interest” so that they are displayed in priority on the interface.

**Figure 2:**
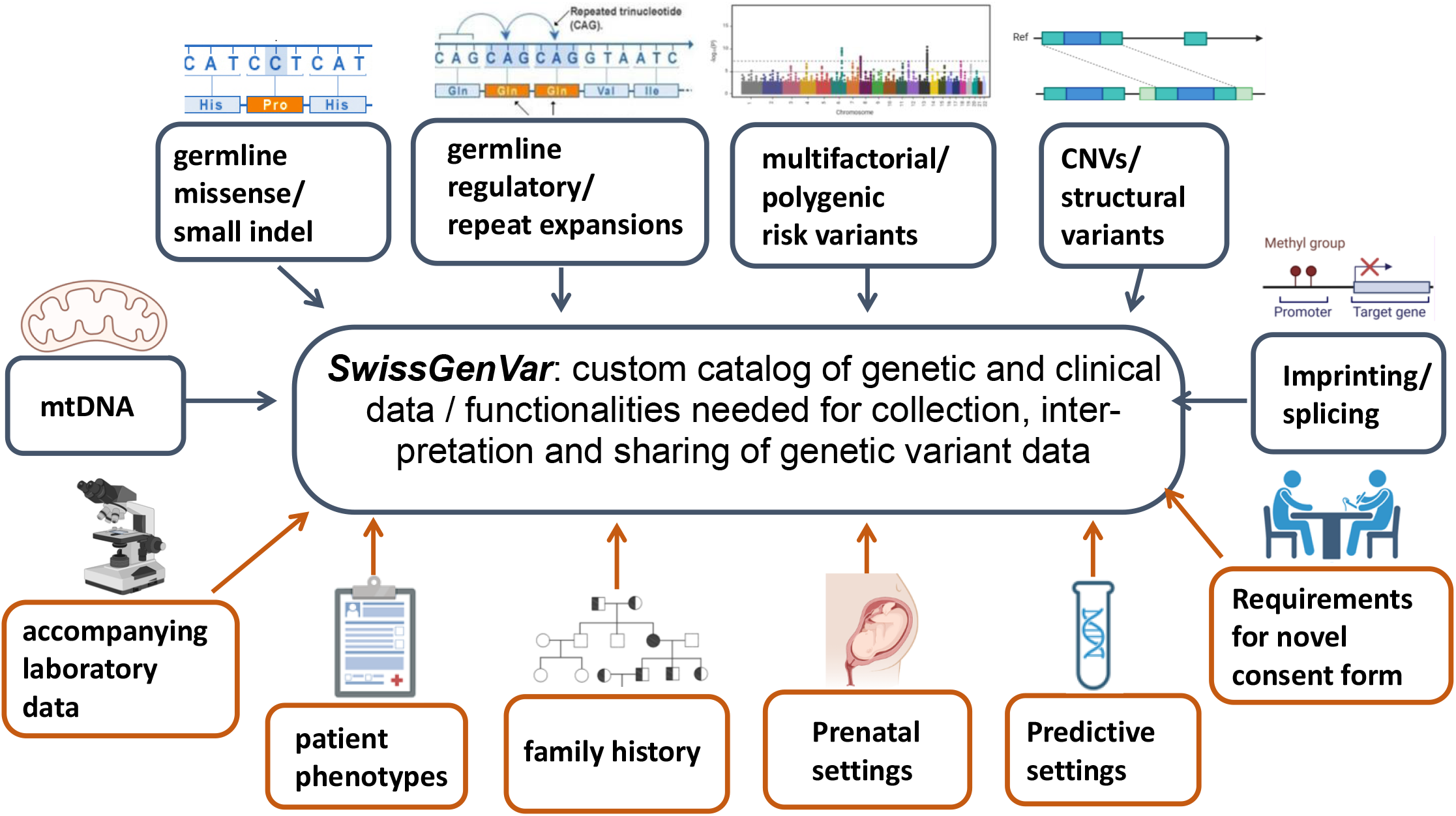
Synopsis of the clinical (*orange*) and laboratory (*grey*) working groups. These elaborated minimal and extended data sets of genetic and clinical data as well as functionalities pertinent to the collection, sharing, and interpretation of genetic variants. At the operational level, a regularly meeting cross-expert team was installed for content implementation.

Additionally, using a local instance of the Ensembl Variant Effect Predictor (VEP), *SwissGenVar* automatically retrieves publicly available annotations for each variant like gnomAD (Genome Aggregation Database) population frequency, variant effect, and the presence of the variant in public databases such as NCBI ClinVar. The implementation of additional public annotations by the integration of APIs from further data sources is being investigated. For the patients’ phenotypic features, *SwissGenVar* allows clinical experts to manually enter clinical information and specific findings relevant to the variant assessment on their patients *via* its web interface using standardized vocabularies agreed upon during the project. Only the data providers are allowed to modify their own data in case corrections or clinical data are added.

### 3.4 *SwissGenVar* database structure, data query, and data display

We developed a graphical user interface to visualize and query the data, enabling the users to explore genetic variants in a gene and/or patient of interest or to retrieve patients with specific phenotypic features. Queries can be issued either from a variant or a patient query page (Figure 3). This interface allows the creation of a custom query based on the user’s interest, with one or multiple criteria filters to search the database and display all the variants or patients matching the selected filtering criteria. The query result is displayed in a variant or patient results table, respectively, that show only selected comparable/searchable information items. However, once a specific variant or patient is selected by clicking on the corresponding row of a results table, the user can access the individual detailed page providing more granular information about the variant or the patient of interest. Thus, the detailed variant page comprises a table of all the patients harboring this specific variant along with selected related information as well as shows automatically retrieved variant annotations as detailed in Tab. 1. The detailed patient page contains a table of variants detected in the patient of interest (obtained from the VCF files) and provides diverse phenotypic features. When no filter is used, the variant and patient tables list all the variants of interest by default and patients present in the database. The pages “Uploaded patients” and “Transferred VCF files”, which are accessible *via* the “My Data” selection panel or menu at the top of the interface, support the users in the management of their own data and provide an overview of their submitted patients and transferred VCF files including their (validation) status. Under the detailed page of the individual patients, the clinical partner of the submitting institution can complement the patient entry with a standardized data set of non-identifying clinical and demographical information and add the granular history of medical contacts with the clinical/phenotypic findings obtained and potential genetic diagnoses. Additionally, the data provider can prioritize clinically (potentially) significant variants by flagging them as of interest (by clicking on the star symbol on the left in the variant table), which is likewise possible directly on the detailed page of the respective transferred VCF file. Finally, the application provides the option of adding variants and patients to a user’s favorite list under the individual detailed page. A notification system will be established to inform the users about any changes or updates concerning their variants or patients of interest.

**Figure 3.**
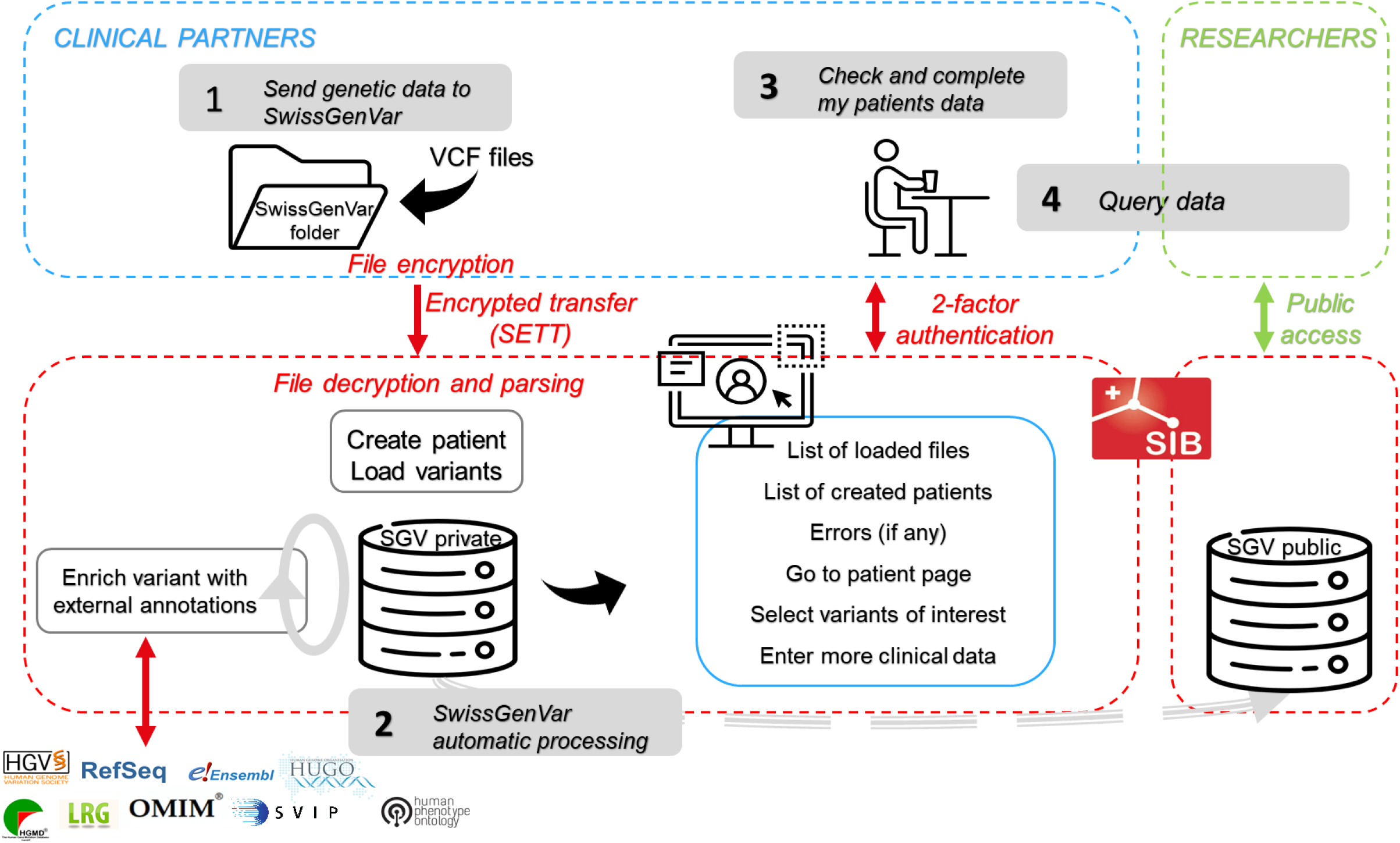
*SwissGenVar* application workflow. The *SwissGenVar* partners provide the genetic variant data to the *SwissGenVar* database in VCF format. The data are encrypted and securely transferred (*step 1*) using the SPHN Secure Encryption and Transfer Tool (SETT). Upon transfer to the *SwissGenVar* private/main application server, the files are decrypted and parsed to create patient entries and load the genetic variants into the platform. The variant entries are automatically enriched with selected external public annotations (*step 2*). At this stage, the partners can connect to their protected account using two-factor authentication to check the transfer of their data files and start adding clinical information on their patients directly on the *SwissGenVar* interface (*step 3*). They can also query the full database to go to specific patient pages and select variants of interest using multiple pre-defined filters (*step 4*). In a future step, *SwissGenVar* will also integrate a publicly accessible platform of aggregated variant-related and clinical information for personalized medicine research.

**Figure 4.**
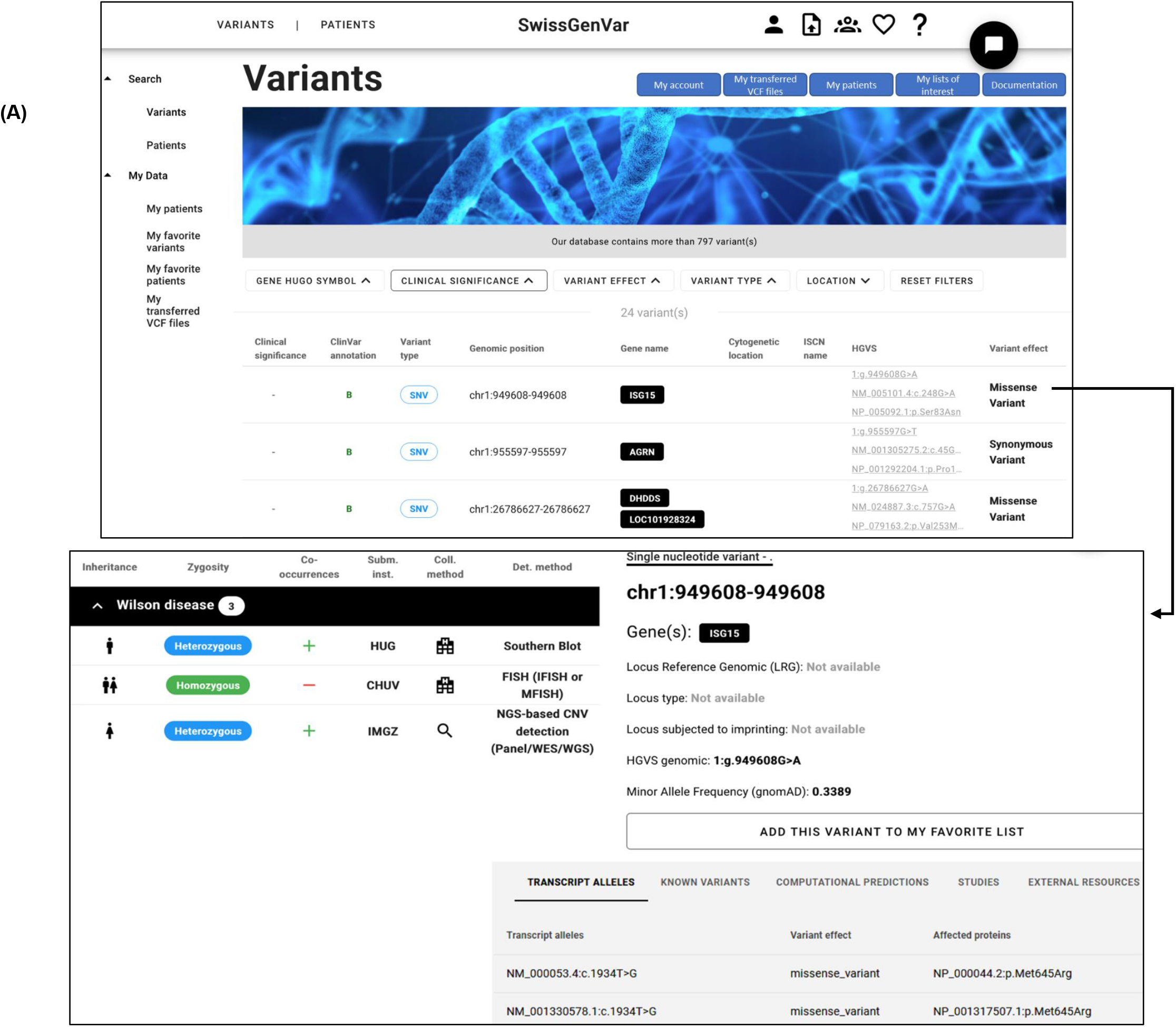
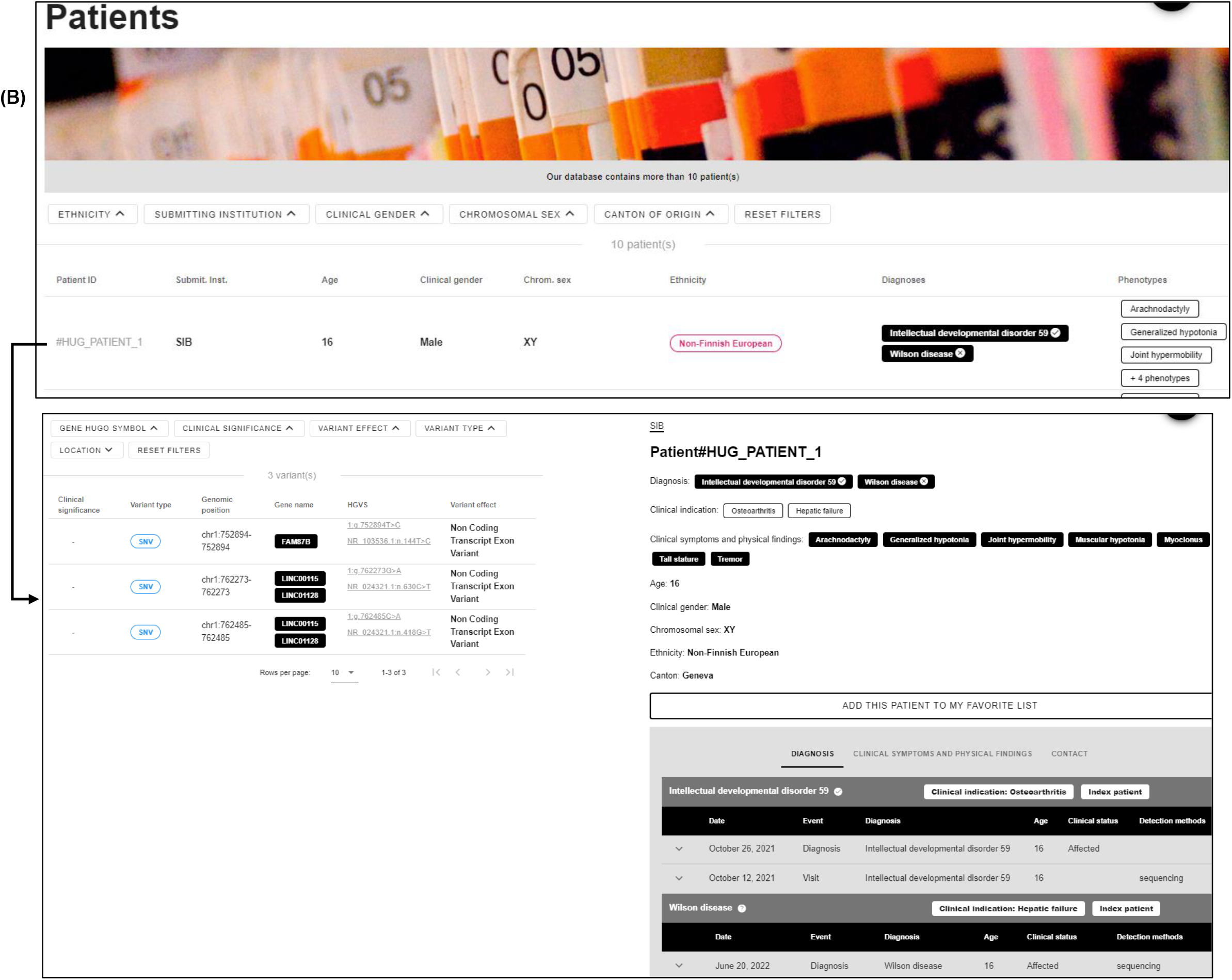
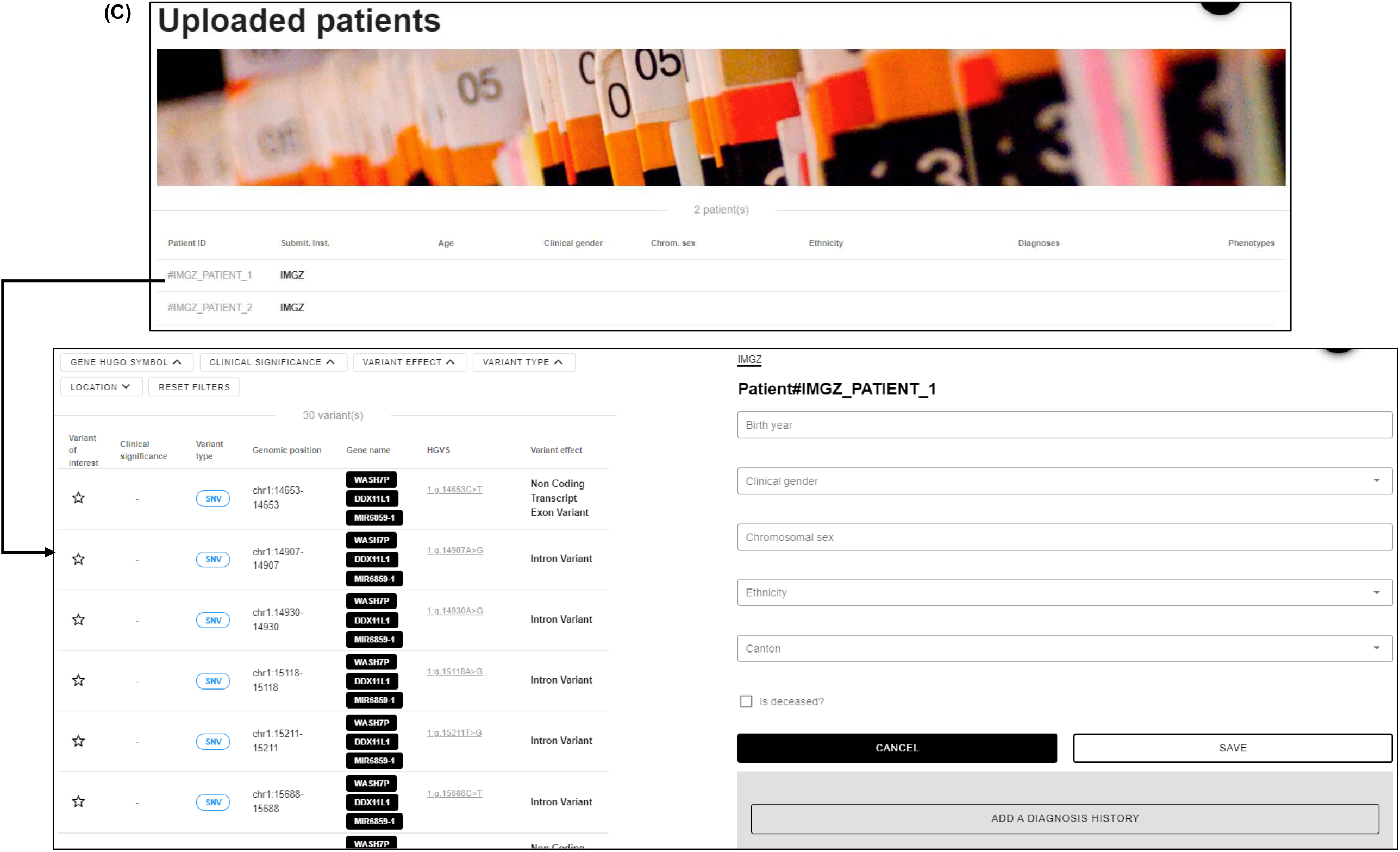
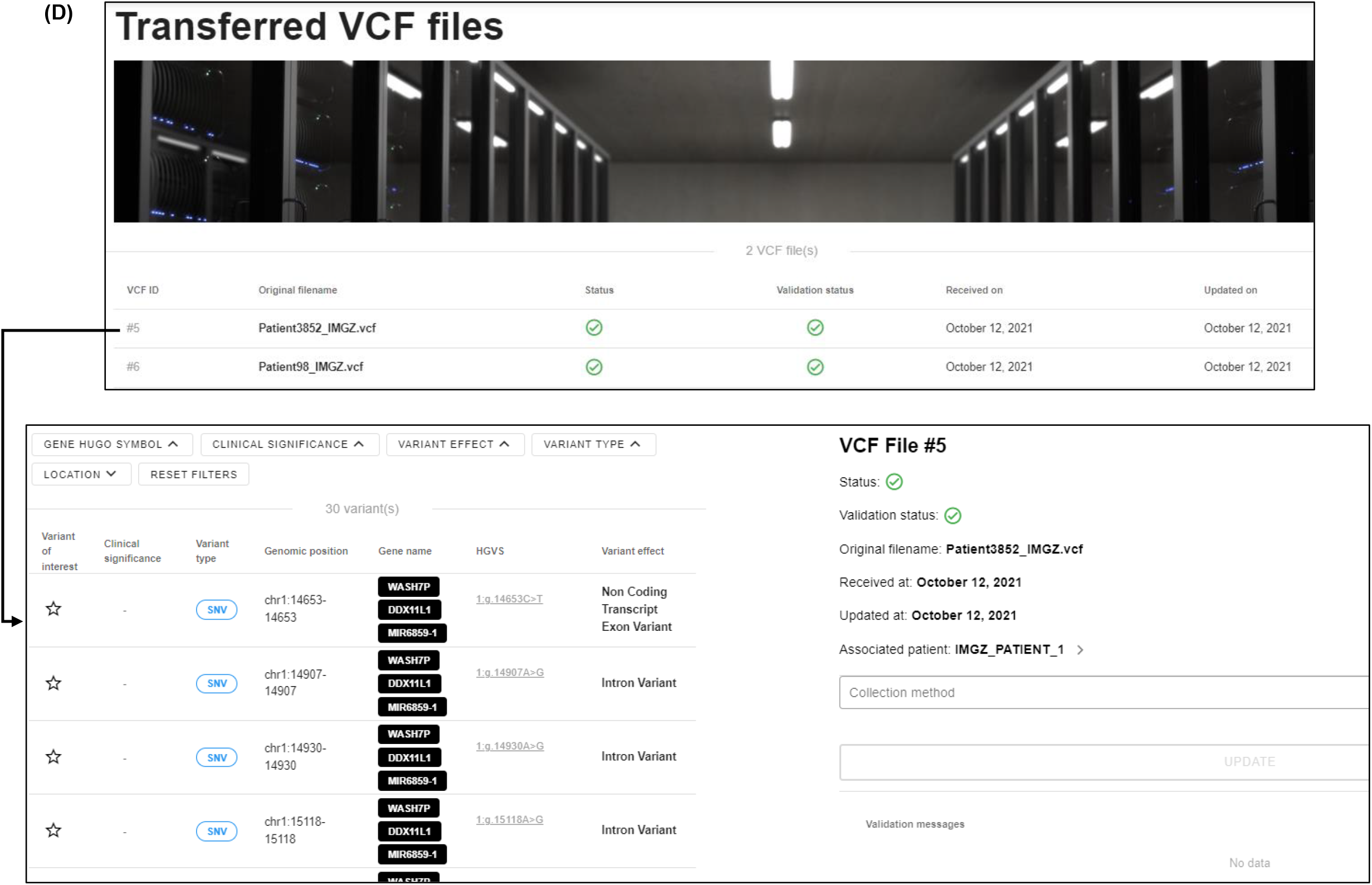
*SwissGenVar* variant (**A**) and patient (**B**) query pages. These pages consist of the filters bar (*1*); table of all/matching variants/patients with selected annotations (*2*); search panel to switch between variant and patient query page (*3*); selection panel “My data” (*4*) to view and edit the patients submitted by the user (under “Uploaded patients” overview and the detailed page (**C**)), respectively) to view the personal lists of the variants/patients of interest, or to view and edit the VCF files transferred by the user (under “Transferred VCF files” overview and the detailed page (**D**), respectively); menu bar (*5*) with a personal account and link to VCF upload service. By clicking on a row of the respective table, the users will be redirected to the individual detailed page providing further information, and in the case of the “My patients” and “Transferred VCF files” pages, to edit their own patients and VCF files. Of note, all presented data are for fictious individuals, not real patients.

## 4. Discussion

*SwissGenVar* aims to use datasets with general consent or with dedicated *SwissGenVar* consent to evaluate the landscape of (clinically relevant) genetic variants in Switzerland to improve variant interpretation and risk assessment by studying genotype-phenotype relation and the natural history of genetic predispositions and disorders. This shall increase our knowledge and result in respective standard operating procedures and structures for improved patient care. Therefore, *SwissGenVar* intends to be a nationwide repository for genetic findings in available and consented genetic data sets across all five academic Medical Genetic institutions in Switzerland and to jointly assess their clinical significance to implement standard operating procedures and improved genetic diagnostics and patient care. As main achievement, *SwissGenVar* allows for the collection and sharing of genetic and associated clinical data *via* a secure data transfer and access/query by the project partners. At the same time, it provides a platform for knowledge sharing about variant-related evidence to harmonize and upscale their significance interpretation at clinical grade with interoperability with international efforts.

For this purpose, *SwissGenVar* supports granular multifactorial filtering for variants and patients in separate query interfaces and details “in-house” variant-related and clinical evidence such as data from local mutation and clinical databases, and segregation as well as experimental analyses. Additionally, *SwissGenVar* enables its users to submit published information like published literature reports and functional studies as well as includes publicly retrievable variant annotations and links to well-established variant databases following international standards. Compared to existing genotype-phenotype/variant databases, the integration of the complete set of variant calls from the transferred VCF files and of the granular history of medical contacts and portfolio of phenotypic findings can be regarded as a big advantage in addition to the collection of genetic variants found in Swiss subpopulations [45, 46]. This allows for the comprehensive clinical assessment of variants in the synopsis of co-occurring candidate variants and the respective clinical features of the variant-carrying individual, which is supported by the option to flag several variants as of interest in the corresponding VCF files. To prompt expert discussions about significance interpretation, a notification system will inform users about any changes or updates concerning the classification of individual variants or patients of interest. Finally, the *SwissGenVar* project has strongly contributed to harmonizing diagnostic practices among the participating institutions by defining and standardizing ontologies for variant and related clinical data. The ontology catalogue was provided to the SPHN Data Coordination Center (DCC) [47] to serve as a basis for other (and follow-up) projects in medical genetics.

Individual findings may be followed up and, depending on the consent provided, clearly pathogenic findings with high predictive value may be fed back to the referring medical geneticist for genetic counseling of the patient. The knowledge gained for individual variants shall be annotated in the *SwissGenVar* database and may become publicly accessible in a public outlet of the platform integrating interfaces with international database efforts. So far, the platform is available to partner groups only, but with a scope for expansion to further academic and non-academic institutions.

## 5. Conclusions

In conclusion, *SwissGenVar* provides a protected platform for the nationwide collection of genetic germline variants and the sharing of related evidence and curated variant significance interpretations by clinical genetic experts, integrating a consistent genetic variant file upload and semi-automated annotation/curation pipeline. Thus, *SwissGenVar* may be regarded as a necessary first step to harmonize and scale-up clinical-grade genetic testing in Switzerland, thereby fostering personalized health research involving genetic risk stratification and disease classifications.

## Data Availability

The SwissGenVar documentation and public website are available at https://pages.sib.swiss/project/swissgenvar-doc/ and https://pages.sib.swiss/project/swissgenvar/, respectively. The project GitLab site is available at URL https://gitlab.sib.swiss/clinbio/swissgenvar/sgv-knowledge/-/tree/master.

## Author Contributions

Conceptualization: M.A., W.B., S.C., A.S., A.S.F., V.B. and A.R.; Methodology: D.K., D.T., L.F., I.F., P.J., S.K., F.M., S.M., T.N., J.S., C.Z., M.A., W.B., S.C., A.S., A.S.F., V.B. and A.R.; Software: D.T. and V.B.; Data Curation: D.K., D.T., L.F., I.F., P.J., S.K., F.M., S.M., T.N., J.S., C.Z., M.A., W.B., S.C., A.S., A.S.F., V.B., A.R.; Writing – Original Draft Preparation, D.K., D.T., V.B. and A.R.; Writing – Review & Editing: D.K., D.T., L.F., I.F., P.J., S.K., F.M., S.M., T.N., J.S., C.Z., M.A., W.B., S.C., A.S., A.S.F., V.B., A.R.; Visualization: D.K., D.T., V.B. and A.R.; Project Administration: A.R.; Funding Acquisition: A.R.

## Institutional Review Board Statement

Not applicable.

## Informed Consent Statement

Not applicable. All presented data are for fictious individuals, not real patients.

## Data Availability Statement

The *SwissGenVar* documentation and public website are available at https://pages.sib.swiss/project/swissgenvar-doc/ and https://pages.sib.swiss/project/swissgenvar/, respectively. The project GitLab site is available at URL https://gitlab.sib.swiss/clinbio/swissgenvar/sgv-knowledge/-/tree/master.

